# A Digital Protein Microarray for COVID-19 Cytokine Storm Monitoring

**DOI:** 10.1101/2020.06.15.20131870

**Authors:** Yujing Song, Yuxuan Ye, Shiuan-Haur Su, Andrew Stephens, Tao Cai, Meng-Ting Chung, Meilan Han, Michael W. Newstead, David Frame, Benjamin H. Singer, Katsuo Kurabayashi

## Abstract

Despite widespread concern for cytokine storms leading to severe morbidity in COVID-19, rapid cytokine assays are not routinely available for monitoring critically ill patients. We report the clinical application of a machine learning-based digital protein microarray platform for rapid multiplex quantification of cytokines from critically ill COVID-19 patients admitted to the intensive care unit (ICU) at the University of Michigan Hospital. The platform comprises two low-cost modules: (i) a semi-automated fluidic dispensing/mixing module that can be operated inside a biosafety cabinet to minimize the exposure of technician to the virus infection and (ii) a 12-12-15 inch compact fluorescence optical scanner for the potential near-bedside readout. The platform enabled daily cytokine analysis in clinical practice with high sensitivity (<0.4pg/mL), inter-assay repeatability (∼10% CV), and near-real-time operation with a 10min assay incubation. A cytokine profiling test with the platform allowed us to observe clear interleukin −6 (IL-6) elevations after receiving tocilizumab (IL-6 inhibitor) while significant cytokine profile variability exists across all critically ill COVID-19 patients and to discover a weak correlation between IL-6 to clinical biomarkers, such as Ferritin and CRP. Our data revealed large subject-to-subject variability in a patient’s response to anti-inflammatory treatment for COVID-19, reaffirming the need for a personalized strategy guided by rapid cytokine assays.

## Introduction

With the global outbreak of the novel coronavirus pneumonia (COVID-19), accumulating evidence^1-3^ indicates that cytokine storm or cytokine release syndrome (CRS) is associated with severe illness. CRS is observed in several disease states associated with dysregulated immunity, including as a consequence of CAR-T cell immunotherapy^4^, a manifestation of hemophagocytic lymphohistiocytosis (HLH) in malignancy, macrophage activation syndrome in autoimmune disease^5^, or severe sepsis^6^. Selective cytokine blockade is a mainstay of care for CRS related cancer immunotherapy^4, 7, 8^, and macrophage activation syndrome^9^. In COVID-19, early translational studies suggest that high serum cytokines are a result of a complex interplay between lymphocytes and myeloid cells^10^. Modulation of cytokine signaling pathways is currently the subject of over 50 clinical trials worldwide^11^. However, most studies enroll based on clinical criteria without rapid assessment of specific cytokine levels, despite delivering therapies that are targeted to specific cytokines, such as interleukin (IL)-6. In our center, the current clinical practice is to use a variety of less specific surrogate markers, such as ferritin and CRP, to gauge a patient’s overall level of inflammation. While cytokine levels are being checked in patients with severe COVID-19, in practice, the results of these tests return in days, not hours. Ideally, treating physicians would understand the “real-time” level of a variety of cytokines in a particular patient before administering specific medications to blunt cytokine storm in critical illness, which urgently requires a low-cost near-bedside multiplex cytokine profiling assay with a rapid assay turnaround.

Digital immunoassay ^12, 13^ has been considered as the next generation protein detection method which provides single-molecular sensitivity (aM-fM) detection by digitizing and amplifying enzymatic reaction in extremely confined volumes (fL-nL). Several groups invented microfluidic platforms for lab-on-a-chip operation of digital assays ^14-17^ and notably, Yelleswarapu et al ^18^ demonstrated a mobile-phone-based, droplet microfluidic digital immunoassay for point-of-care (POC) settings. However, few studies have implemented a digital assay platform applicable to the clinical treatment of a COVID-19-induced cytokine storm. If continuous monitoring of the cytokine profiles of a COVID-19 patient is needed, the assay requires more than speed, sensitivity, and multiplex capacity. Other important but often overlooked requirements include (1) flexibility of running a small number of samples based on the demand of the physician with minimum preparation; (2) great inter-assay precision between multi-time point measurements, which is not an issue in conventional large batch-based retrospective tests; (3) a low-cost, compact, automated fluidic handling and readout instrumentation that can be operated inside the bio-safety cabinet with minimum user exposure to virus-contaminated blood samples.

Here, we report the development and application of an automated digital assay platform using a method termed the “pre-equilibrium digital enzyme-linked immunosorbent assay (PEdELISA) microarray” for rapid multiplex monitoring of cytokine: IL-6, TNF-α, IL-1β and IL-10 from COVID-19 patients admitted to the ICU in the University of Michigan hospital. The PEdELISA microarray analysis employs magnetic beads trapped into spatially registered microwell patterns on a microfluidic chip. The locations of the microwell patterns on the chip indicate which target analytes are detected. Unlike the existing digital assays, our method employs an approach of quenching the assay reaction entirely on-chip at an early pre-equilibrium state. This approach achieves near-real-time assay speed (<10 min incubation) with a clinically relevant fM-nM dynamic range without losing assay linearity. Furthermore, using a simple microfluidic spatial encoding technique and machine learning-based image processing algorithm, we achieved multiplex detection with high-accuracy counting and eliminated significant bead loss faced by the commercial state of the art platform^19^. The advancements of our digital assay demonstrated here enable it as a great candidate for near-bedside cytokine profiling with the combination of speed and sensitivity, both greater than those of current analog^20-23^ and label-free POC diagnostic systems^24-27^.

## Results and Discussion

The PEdELISA microarray assay platform comprises a cartridge holding a disposable microfluidic chip with capture antibody (CapAb)-conjugated magnetic beads pre-settled in the designated microarray locations according to the antibody type, a parallel pipetting module controlled by Arduino for on-chip fluidic dispensing and mixing, and a 2-axis cartridge scanning and fluorescence imaging module (Fig. 1A, see Supporting Information for system details). In this setup, the disposable microfluidic cartridge (Fig. 1A, inset) was designed to handle 16 samples per chip with up to 16-plex maximum capacity. The chip contains two polymethyl methacrylate (PMMA) layers top (venting) and bottom layer (substrate) with countersink connectors that are seamlessly interfaced with fluidic dispensing tips, a thin polydimethylsiloxane (PDMS) layer (200 µm) which contains fL-sized microwell arrays for digital assay, and a polyethylene terephthalate (PET) thin (120 µm) film with microfluidic channels fabricated by laser cutting (see Fig. S1 for cartridge fabrication). The use of the materials and processing methods significantly reduced the chip manufacturing cost (< $0.5/chip).

**Figure 1.**
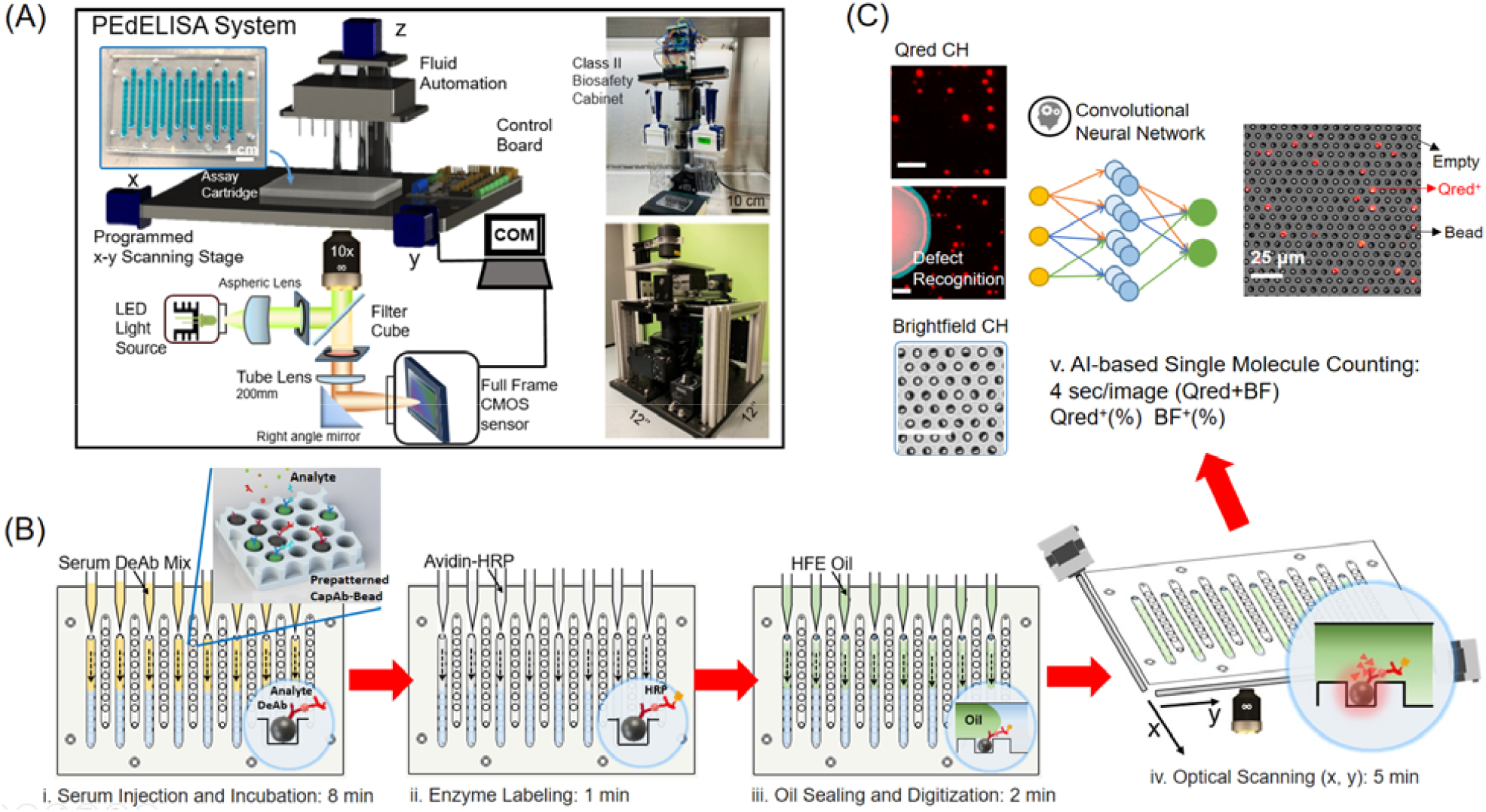
PEdELISA microarray assay platform for COVID-19 patient cytokine storm profiling. (A) Schematic and photo image of the assay system in a biosafety cabinet. The platform comprises a cartridge holding a disposable microfluidic chip (inset), an automated fluidic dispensing and mixing module, and a 2D inverted fluorescence scanning module. (B) The 4-step assay procedure includes (i) automated injection and subsequent on-chip mixing of serum and a detection antibody solution with capture antibody-coated magnetic beads pre-deposited in microwell arrays, which is accompanied by a short incubation (8-min) and followed by washing (2-min), (ii) HRP enzyme labeling (1-min), followed by washing (5-min), (iii) fluorescence substrate loading and oil sealing (2-min), and (iv) x-y optical scanning and imaging (12-min). (C) Data analysis using a convolutional neural network-guided image processing algorithm for high throughput and accurate single-molecule counting that corrects image defects and accounts for signal intensity variations. Both the fluorescence substrate channel (Qred CH) and brightfield channel (BF CH) are analyzed to calculate the average number of immune-complexes formed on each bead surface. The unlabeled scale bars are 25 μm.

The PEdELISA assay was carried out by the programmed pipetting module that allowed for microfluidic loading and handling in a consistent and repeatable manner (Fig. S2A). The module first mixed patient samples or assay standards with a detection antibody (DeAb) solution and then loaded them into the cartridge in parallel, followed by 50 automated cycles of on-chip mixing during incubation (8 min), washing (2 min) and enzyme labeling (1 min), washing (5 min), substrate loading, and oil sealing (Fig. 1B, see Supporting Information for assay details). The chip was subsequently scanned and imaged by the compact and low-cost (<$5000) fluorescence imaging module using a consumer-grade CMOS camera (Fig. S2B), and the data was analyzed by a high-throughput in-house image processing algorithm based on convolution neural network and parallel computing (Fig. 1C). This algorithm performed autonomous classification and segmentation of image features such as microwells, beads, defects, and backgrounds, so that the digital assay counting results were generated without human supervision. The assay involved some minor manual work for assay reagent preparation and serial dilution, fluid waste collection, z-axis focusing, and origin/endpoint positioning to trigger the optical scanning.

We ensured the x-y optical scanning motion control accuracy each time by repetitively scanning and imaging the microarray structures on the cartridge. Post-image processing was used to calculate the x, y offset, which may be induced by the imperfection of system alignment, lead screw backlash, or motor step missing. We developed a mathematical algorithm to correct these offsets, and the scanning module was able to achieve less than 5 µm bidirectional repeatability and 0.31 µm minimum incremental movement (Fig. S3). Using the programmed fluidic dispensing system, we optimized the assay reaction parameters (incubation time and reagent concentration) and achieved a limit of detection (LOD) less than 0.4 pg/mL with both assay reaction and labeling incubation time in 9 min (Table 1). We also assessed the 4-plex assay’s specificity and signal to noise ratio (SNR) by spiking-in each cytokine analyte in 100% fetal bovine serum buffer (FBS) to mimic the patient serum detection. Fig 2A shows the assay results of “all-spike-in,” “single-spike-in,” and “no-spike-in” using 200 pg/mL recombinant cytokine standards (a typical clinical threshold for cytokine storm). Negligible antibody cross-reactivity was observed between each cytokine analyte and SNR=488.0 was calculated on average (averaged assay signal over background signal).

**Table 1.** Limit of detection (LOD), limit of Quantification (LOQ), and coefficient of variation (CV) of PEdELISA for a panel of 4 cytokines. Here, the LOD and LOQ values were determined from the blank signal + 3σ and the blank signal + 10σ, respectively. The intra-assay CV was determined by quadruplicate measurements of five COVID-19 patient samples at the range of 6-600 pg/mL in both near-real-time and retrospective assay modes. The inter-assay CV was determined by taking the root-mean-square average of signals from 40, 200, and 1000 pg/mL assay standard in 10-day continuous measurements of COVID-19 patients.

| Cytokine Type | Assay Blank<br>(Average molecule per bead) | Assay Blank+$3\sigma$<br>(Average molecule per bead) | LOD<br>(pg/mL) | LOQ<br>(pg/mL) | Intra-Assay CV (%) | Inter-Assay CV (%) |
| --- | --- | --- | --- | --- | --- | --- |
| IL-1β | 0.000143 | 0.000347 | 0.191 | 1.188 | 4.68 | 9.98 |
| TNF-α | 0.000179 | 0.000379 | 0.198 | 1.889 | 8.77 | 11.66 |
| IL-10 | 0.000136 | 0.000378 | 0.350 | 1.552 | 5.70 | 9.63 |
| IL-6 | 0.000189 | 0.000309 | 0.377 | 2.378 | 4.55 | 10.80 |

**Figure 2.**
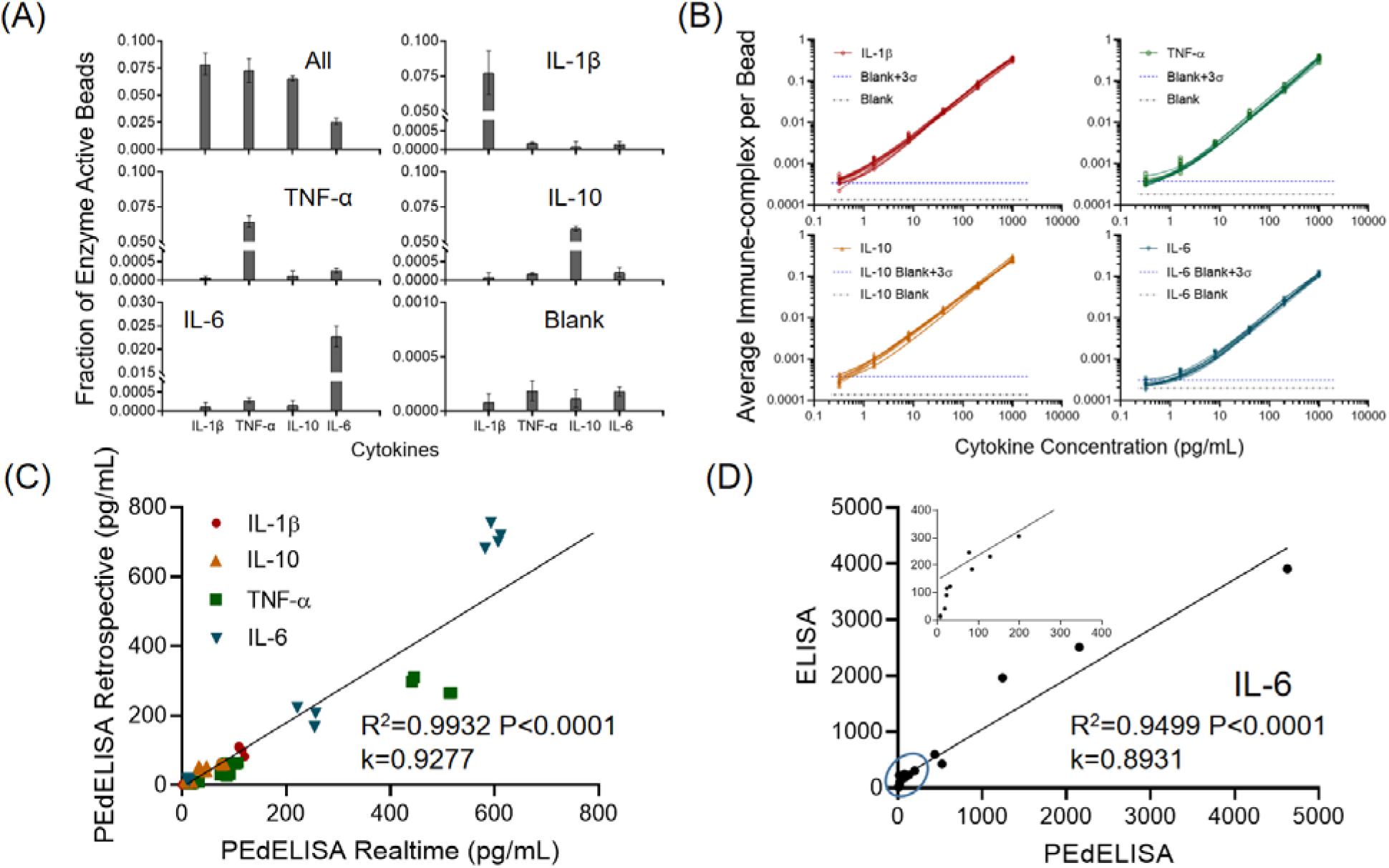
PEdELISA assay characterization. (A) Assay specificity test with “all-spike-in,” “single-spike-in,” and “no-spike-in” (negative) of recombinant cytokine standard at 200 pg/mL in fetal bovine serum (FBS) buffer. (B) Daily COVID-19 patient assay standard curves for four cytokines from 0.32 pg/mL to 1000 pg/mL in FBS (10 curves for each cytokine obtained over 10 workdays). The data points were fitted with four-parameter logistic (4PL) curves. The black dotted line represents the signal level from a blank solution. The blue dotted line shows 3σ above the blank signal, which is used to estimate the limit of detection (LOD) for each cytokine. (C) Linear correlation (R^2^=0.99, P<0.0001) between rapid measurements of fresh samples and retrospective measurements of stored samples (1 freeze-and-thaw at −80 °C) in quadruplicate for 5 representative COVID-19 patients. (D) Good agreement (R^2^=0.95, P<0.0001) observed between single-plex IL-6 ELISA and multiplex PEdELISA measurements for 16 COVID-19 patients. The inset shows the circled region.

In order to facilitate the care of patients with COVID-19 at the University of Michigan Hospital, we undertook a pragmatic study to rapidly return same-day cytokine levels to the clinical teams treating critically ill COVID-19 patients in ICU at the physicians’ request from April 9^th^ to May 29^th^ in 2020. Given the investigational nature of the assay, patients or their representatives provided informed consent for cytokine measurements to be provided for clinical use (UM IRB HUM00179668). To ensure the accuracy of our data, the COVID-19 patient samples were run in quadruplicate with an assay standard curve calibrated every day. Fig. 2B shows assay standard curves that were accumulated in 10 different workdays of the patient cytokine monitoring period. The multiple assay standard curves yielded excellent repeatability with the inter-assay coefficient of variation (CV) of ∼ 10% due to the programmed fluidic handling and reaction (Table 1). We also characterized the intra-assay CV for five representative COVID-19 patient serum samples with cytokines at concentrations ranging from 6-600 pg/mL, each tested in quadruplicate measurements (Table 1). We compared assay data for these five patients resulting from near-real-time measurements of fresh samples drawn daily and retrospective measurements of stored samples after one freeze-thaw cycle. We observed a good linear correlation (R^2^=0.99) between the two measurement modes except for TNF-α. This suggests that TNF-α in the stored serum could degrade by 20-40% after the freeze-and-thaw banking at −80 °C (Fig. 2C). Additionally, to validate our PEdELISA microarray assay, we compared the assay results with those of a conventional single-plex ELISA method that retrospectively measured 15 banked samples from identical patients. Because conventional ELISA requires a much larger sample volume (>200 μL for each measurement, in duplicate per analyte) relative to PEdELISA, it was practically difficult for us to manage the acquisition of a sufficiently large blood sample volume from critically ill COVID-19 patients. Therefore, we only validated our assay against IL-6 detection results (Fig. 2D). The data between these two methods overall matched linearly (R^2^=0.95, P<0.0001). Some discrepancy was observed at concentrations below 50 pg/mL and may be potentially due to the limited sensitivity and linearity of the ELISA assay (Fig. 2D inset).

Fig. 3A shows a typical timeline of our daily cytokine profile measurement completed within 4 hours after the blood draws in the ICU. The assay itself could be performed with a sample-to-answer time as short as 30 min for typical non-COVID-19 serum samples. However, in the practical operation of our test, a larger amount of time was spent on sample processing, transport, and team coordination, as well as biosafety and disinfection protocols in handling COVID-19 samples. Nevertheless, the <4-hour blood draw to result turnaround is still rapid as compared to typical clinically deployed tests. Our rapid cytokine measurement in patients with respiratory failure due to COVID-19 revealed significant subject-to-subject heterogeneity despite all patients being critically ill. As expected, interruption of IL-6/IL-6R signaling in patients who received tocilizumab resulted in marked elevation of IL-6 levels in the setting of ongoing illness (p<0.0001, Fig. 3B).^28^ Among patients who did not receive tocilizumab, we observed a large degree of variability in IL-6 levels, with a quarter of subjects having IL-6 <15 pg/mL, the median value of 106 pg/m, and the CV of 114%. Variability of TNF-α (CV 164%) and IL-1β (CV 193%) was driven by a small number of subjects with elevated levels. However, like IL-6, levels of IL-10 were also broadly distributed in patients who had not received tocilizumab (CV 93%).

**Figure 3.**
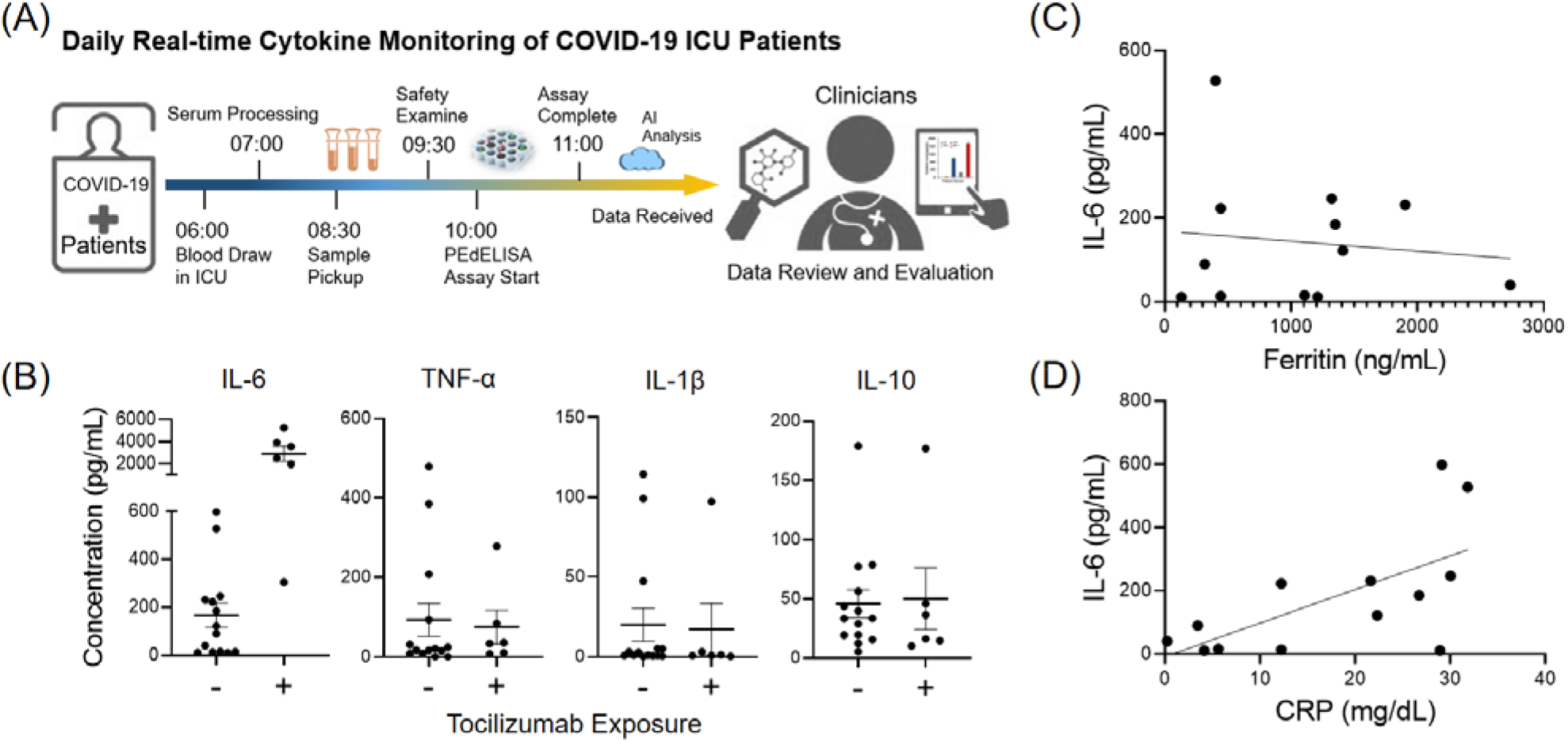
(A) Timeline of daily near-real-time COVID-19 cytokine measurement. (B) Statistical group analysis of patients that are dosed/undosed with Tocilizumab. Significant elevations of IL-6 levels were observed after the treatment of Tocilizumab (P<0.0001). (C)-(D) Correlation of cytokine IL-6 to Ferritin and C-Reactive Protein (CRP), standard clinical inflammatory biomarkers. Ferritin does not correlate well with IL-6 (R^2^ = 0.01, P=0.71). CRP correlates with IL-6 (R^2^ =0.41, P=0.018) better, but the IL-6 levels were widely distributed for patients with high levels of CRP.

Given the heterogeneity of cytokine levels in critically ill patients with COVID-19, we asked whether IL-6 levels were reflected in surrogate biomarkers. In current, rapidly evolving clinical practice, the presence of cytokine storm and risk of clinical deterioration is frequently judged by inflammatory markers such as CRP and ferritin in the absence of direct cytokine measurement. Ferritin did not predict IL-6 levels (Fig. 3C, R^2^ = 0.01, P=0.71). CRP was significantly associated with IL-6 (Fig. 3D, R^2^ =0.41, P=0.018). However, this association was driven by low IL-6 in subjects with low levels of CRP, while IL-6 values in subjects with high CRP were widely distributed. Neither CRP nor ferritin is a reliable predictor of IL-6. Note that our pragmatic study of rapid cytokine measurements in patients with COVID-19 was designed to provide information to clinicians, rather than systematically study the biology of COVID-19. We therefore, enrolled subjects without regard to time from the onset of infection. Furthermore, due to these subjects’ critical illness, many received empiric antibiotic therapy, limiting our ability to determine bacterial co-infection. These factors, as well as our small sample size, may have contributed to the heterogeneity of the cytokine response. Nevertheless, all subjects in this study were critically ill and had respiratory failure, underscoring the diversity of biological mechanisms that may lead to critical illness in COVID-19 and the importance of measuring, rather than inferring, cytokine storm.

## Conclusion

Timely intervention of cytokine storm guided by rapid cytokine measurement is critical for the management of severe COVID-19 infections resulting in respiratory failure. To this end, we have developed a microfluidic digital immunoassay platform that enables rapid 4-plex measurement of cytokines in COVID-19 patient serum. Our assay employed single-molecule counting for an antibody sandwich immune-complex formation quenched at an early pre-equilibrium state. The pre-equilibrium approach resulted in a detection limit < 0.4 pg/mL and a linear dynamic range of 10^3^ while requiring a total assay incubation time as short as 10 min. The platform incorporates a programmed fluidic dispensing and mixing module and a compact optical reader module for microfluidic analysis using low-cost disposable chips manufacturable at a large scale. Each chip contains spatially encoded microwell array patterns with capture antibody-coated magnetic beads pre-deposited for multiplex cytokine detection. The 4-plex on-chip measurement with a 15 µL sample volume showed negligible sensor cross-talk. The programmed fluidic handling and mixing module permitted high inter-assay repeatability (∼10% CV). Our assay platform with the combination of high sensitivity, speed, and fidelity allowed us to complete a whole cytokine profiling test from initial blood draw with critically ill COVID-19 patients to data delivery to physicians within 4 hours.

Anti-IL6 therapy in COVID-19 is under investigation in multiple clinical trials and has been employed in clinical practice as an off-label use. Preliminary results from trials of the anti-IL6 receptor antibody sarilumab have not demonstrated efficacy in moderately ill patients and trials continue in critically ill patients^29^. Notably, our results here highlight the heterogeneity of cytokine response even among critically ill COVID-19 patients and the poor ability of surrogate inflammatory markers to predict an IL-6 response. These results confirm that rapid, reliable and repeatable direct cytokine measurement is needed to facilitate precision administration of anti-cytokine therapies only in patients who are experiencing cytokine storm. Our digital immunoassay platform may provide a promising means to enable such a precision medicine strategy in the pharmacotherapeutic management of life-threatening cytokine storm in COVID-19.

## Materials and Methods

### Materials

We purchased human IL-6, TNF-α capture, and biotinylated detection antibody pairs from Invitrogen™, and IL-1β, IL-10 from BioLegend. We purchased the corresponding ELISA kits from R&D Systems (DuoSet®). We obtained Dynabeads, 2.7μm-diameter epoxy-linked superparamagnetic beads, avidin-HRP, QuantaRed™ enhanced chemifluorescent HRP substrate, bovine serum albumin (BSA), TBS StartingBlock T20 blocking buffer, and PBS SuperBlock blocking buffer from Thermo Fisher Scientific. We obtained Phosphate buffered saline (PBS) from Gibco™, Sylgard™ 184 clear polydimethylsiloxane (PDMS) from Dow Corning, and Fluorocarbon oil (Novec™ 7500) from 3M™. The automated PEdELISA system was mainly constructed by a micro-controller (Arduino Uno and MEGA 2560), stepper motors and shields (NEMA 17, 0.9 degree, 46 N □ cm, TB6600 and DM542T motor shields), linear rail guide with ballscrews (5 mm/revolution), standard anodized aluminum profiles, clear acrylic boards and other supporting wheels, connectors and parts purchased from Amazon through various vendors. The optical scanning system mainly consists of a consumer-grade CMOS camera (SONY α6100), 10x Objective lens (Nikon, CFI Plan Achro), tube lens (200 mm), optical filter sets (Chroma), halogen light source, LED light source (560 nm), optical mountings and tubings (mainly from Thorlabs and Edmund Optics).

### Antibody Conjugation to Magnetic Beads

We conjugated human IL-6, TNF-α, IL-1β, IL-10 capture antibodies using the epoxy-linked Dynabeads (2.7 μm) with the capture antibody molecules at a mass ratio of 6 μg (antibody): 1 mg (bead) following the protocols provided by Invitrogen™ (Catalog number: 14311D). The beads were then quenched (for unreacted epoxy groups) and blocked with TBS StartingBlock T20 blocking buffer. We stored the antibody-conjugated magnetic beads at 10 mg beads/mL in PBS (0.05% T20 + 0.1% BSA + 0.01% Sodium Azide) buffer sealed with Parafilm at 4 °C. No significant degradation of the beads was observed within the 3-month usage.

### PEdELISA Cartridge Fabrication and Patterning

The disposable microfluidic cartridge used for PEdELISA assay is plastic-based and fabricated by laser cutting and PDMS molding. It has a transparent sandwich structure for optical imaging as shown in Fig. S1. The top and bottom layers are laser-cut using 3.175 mm (1/8 inch) and 1 mm thin clear polymethyl methacrylate (PMMA) boards which contain through-holes for venting and screw assembly purposes. The microfluidic channels (designed with AutoCAD software) are laser-cut through a 120 µm high definition transparency polyethylene terephthalate (PET) thin film (adopted from standard screen protector) which has a silicone gel layer to create a vacuum for securely sealing to the top acrylic layer without adhesives. The power and speed of the laser cutter are optimized to ensure a high-resolution smooth cut so that resistance difference or bubbles generation can be minimized during the fluidic handling process. The femtoliter-sized microwell (d=3.4 μm) array layer (∼300 μm) was made by polydimethylsiloxane (PDMS) through a standard SU-8 molding. First, we constructed SU-8 molds on oxygen plasma treated silicon wafers by standard photolithography which involved depositing negative photoresist (SU-8 2005 MicroChem) layers at 5000 rpm to form the desired thicknesses 3.8±0.1 µm. Subsequently, a precursor of PDMS was prepared at a 10:1 base-to-curing-agent ratio and deposited onto the SU-8 mold by spin coating (300 rpm) and baking overnight at 60 °C. We then transferred the fully cured PDMS thin film onto the bottom acrylic layer using a modified surface silanization bonding method based on a previous publication^30^. We also drilled 2 mm countersink holes (60°) using a benchtop mini drill press (MicroLux®) on the top venting layer for guiding the multi-pin fluidic dispensing connector. Each layer was thoroughly cleaned through water bath sonication and the PET microchannel layer was carefully attached to the top venting layer for the later bead patterning process.

The PEdELISA bead patterning process involves first attaching the bead settling layer (containing long straight PDMS channels perpendicular to the PET microchannel layer) to the PDMS microwell array layer on the bottom PMMA substrate. Then, we prepared 4 sets of a 25 µL bead solution at the concentration of 1 mg/mL for IL-1β, TNF-α, IL-10, and IL-6 bead respectively. The bead solution was loaded into four different physically separate patterning channels in the bead settling layer. After waiting 5 min for beads settling inside the microwells, we washed the patterning channels with 200 uL PBS-T (0.1% Tween20) to remove the unstrapped beads. At this step, we imaged the microarray under the microscope to ensure that the microwells were filled with the beads at a sufficient rate (typically above 50%). If not, the bead mixture solution was reloaded and washed again. Finally, the bead settling layer was peeled off and replaced with PET microchannel and top venting layer. Four layers of the cartridge were sandwiched together using M2 bolt screws. Note that the bonding between the PET layer and the PDMS layer was not permanent but through pressure-based self-sealing, which can be later easily peel off and replaced. We then slowly primed each sample detection channel with Superblock buffer to passivate the cartridge surface and incubated the whole chip for at least 1 hour before the assay to avoid non-specific protein adsorption. The cartridge was typically prepared in batch and sealed in a moisture-controlled petri-dish at room temperature for up to a week with no significant degradation.

### Programmed PEdELISA Assay and Imaging

The automated pipetting system was programmed to first draw 15 µL of the sample solution (patient serum and assay standard) and mix with 15 µL of detection antibody (DeAb) solution in the 96-well tube rack for 20 cycles (25-sec), and then draw 28 µL of the mixed solution and load them into the PEdELISA cartridge by two steps: Step 1. Load 14 µL of the sample-DeAb mix for channel buffer exchange, delay 10-sec for wiping away the original buffer solution inside the channel (1x PBS solution), Step 2. Load the rest of 14 µL, followed with 50-cycles of on-chip mixing (8-min). Then the system drew in 200 µL of washing buffer (PBS-T 0.1% Tween20) and slowly loaded (micro-stepping) into the chip for washing (2-min). Next, the system drew in 40 µL of the avidin-HRP solution and slowly loaded (micro-stepping) into the chip for enzyme labeling (1-min). The chip was washed again with the PBS-T solution for one cycle (200 µL) and 1x PBS solution for another cycle (200 µL), total to reduce the interference between Tween20 and the chemifluorescent HRP substrate later (total 5-min). Finally, the system drew and loaded 30 μL of the QuantaRed substrate solution and then sealed with 35 μL of fluorinated oil (HFE-7500, 3M) for the digital counting process.

The scanning system was used to scan the image of the bead-filled microwell arrays on the PEdELISA cartridge right after the oil sealing step to detect the enzyme-substrate reaction activity. The imaging stage was pre-programmed to follow the designated path to scan the entire chip (64 microarrays) twice: 1. Scan the QuantaRed channel (545nm/605nm, excitation/emission) 2. Scan the brightfield with the transmission light source on. It typically took around 6 min to scan the entire chip for 16 samples in 4-plex detection.

### Pragmatic study of rapid cytokine measurement in COVID-19

This study was approved by the University of Michigan Institutional Review Board (HUM00179668) and patients or their surrogates provided informed consent for the investigational use of this test. Patients with positive SARS-CoV-2 test via PCR and respiratory failure requiring hospitalization in the intensive care unit for heated high flow oxygen or mechanical ventilation were eligible for enrollment. Subjects were approached at the request of treating teams at any point in their disease course after intensive care unit admission. Due to restrictions on patient contact during the COVID19 pandemic, samples were drawn by the subjects’ nurse with routine clinical labs in a serum separator tube and sent to the clinical specimen processing area, where they were centrifuged, aliquoted, and kept at 4°C until analysis. IL-6, TNF-α, IL-1β, and IL-10 were measured and results were posted to the patient’s chart the same day. Ferritin and C-reactive protein (CRP) measurements made during routine clinical care were recorded if available from a sample within 24 hours of the cytokine measurement.

### Statistics

Experiments with synthetic recombinant proteins were performed daily with 2 on-chip repeats averaged to calculate the patient serum cytokine levels. 10-day standard curves using 10 microfluidic cartridges were accumulated to calculate the inter-assay coefficient of variance. The COVID-19 patient serum samples were performed in quadruplicate and averaged for the near-real-time daily cytokine profile monitoring test. Conventional ELISA test was conducted retrospectively for IL-6 in duplicate for selected banked patient samples. Here, Pearson’s R-value was used to quantify the PEdELISA to ELISA correlations and the t-test was used for group analysis of the Tocilizumab treatment. A p-value of < 0.05 was considered to be statistically significant.

## Data Availability

If requested, the authors will make data available for sharing with qualified parties, so long as such a request does not compromise intellectual property interests, invade subject privacy, betray confidentiality, or precede data duration. Data that are shared will include standards and notations needed to interpret the data, following commonly accepted practices in the field. Data will be shared by mailing a CD or USB memory with the data to the requestor from the authors' administrator.

## Author Contributions

B.H.S. and K.K. conceptualized the clinical hypothesis and initialized the research project. Y.S., S.H.S. and A.S. performed the daily patient measurements. M.W.N. processed the daily serum samples. M.H. coordinated the clinical study and sample collection. Y.S., Y.Y., S.H.S., T.C., and M.T.C. designed, built and tested the automated system. Y.S., B.H.S. analyzed the experimental results and all authors contributed to the preparation of this manuscript.

## Acknowledgements

We thank the College of Engineering, Department of Internal Medicine at the University of Michigan for their emergency approval of this COVID-19 related research. We specifically thank the UM Mechanical Engineering machine shop, Mr. Charles Bradley, Donald Wirkner, and Kent Pruss for the great help offered in designing and machining the automated system during the COVID-19 pandemic period. We acknowledge the funding support from the National Institute of Health K08NS101054 (B.H.S), the National Science Foundation CBET1931905 (K.K.), the University of Michigan COVID-19 Response Innovation Grant (B.H.S and K.K), and the University of Michigan Precision Health Scholars Grant (Y.S.).

